# Community-based assessment of mpox seroprevalence and undocumented exposures in Manitoba and Alberta, Canada

**DOI:** 10.64898/2026.09.17.26363367

**Authors:** Elise Gork, Mikayla Hunter, Neve Battle, Nicole Wilson, Samantha Polege, Chelseay Robles, Venice Vincent, Garret Meyer, Andrea Visnjevac, Hannah L. Wallace, Lynora Saxinger, Jared Bullard, Megan Halbrook, Sydney Merritt, Nicole A. Hoff, Ryan S. Noyce, Donald C. Vinh, Rusty Souleymanov, Souradet Shaw, Placide Mbala-Kingebeni, Anne W. Rimoin, Vanessa Meier-Stephenson, Jason Kindrachuk

## Abstract

During the 2022 global mpox outbreak, gay, bisexual and other men who have sex with men (gbMSM) were disproportionately affected. However, the true number of cases is likely higher than reported due in part to the stigma associated with mpox. This stigma may have played an even more influential role on health-seeking behaviour in politically conservative regions, such as the Canadian Prairies. The shifting epidemiology may have also led to misdiagnoses during initial months of the outbreaks. Seroprevalence studies can help assess these gaps in surveillance and population-level immunity. To address this, we conducted a cross-sectional seroprevalence study among 275 individuals who identified as potentially eligible to receive the mpox vaccine in the Canadian provinces of Manitoba and Alberta from June 2024 to December 2025 based on provincial vaccination criteria. This eligibility included, but was not exclusively limited to, gbMSM, gender diverse individuals, and sex workers. Dried blood spots were analyzed using the Meso Scale Discovery *Orthopoxvirus* assay targeting five orthologous mpox virus and vaccinia virus antigens. In total, 14.5% of individuals self-reported receiving the mpox vaccine and of those without reported *Orthopoxvirus* vaccination, 5.9% were seroreactive to multiple mpox virus antigens. These results suggest the potential for undetected mpox circulation within Manitoba and Alberta. While primarily men who have sex with men were eligible for the mpox vaccine, an expansive range of gender and sexual orientation identities were classified as seroreactive, suggesting a misalignment between those who were seroreactive and vaccine eligibility criteria. This emphasizes how broader mpox vaccine eligibility, not restricted by specific gender and sexuality requirements, may better encompass those at risk of infection.

## Introduction

Since the eradication of smallpox, mpox virus (also known as monkeypox virus; MPXV), the etiologic agent of mpox, has emerged as the most prominent *Orthopoxvirus* capable of global expansion. Historically endemic to forested regions of West and Central Africa, MPXV infections cause characteristic lesions to appear on individuals, most commonly on the trunk of the body and extremities. The first clinically recognized case of mpox occurred in 1970 in the Democratic Republic of the Congo (DRC) and succeeding sporadic cases occurred via zoonotic transmission (Breman et al., 1980; Ladnyj et al., 1972). From the 1980s to 2007, a dramatic 20-fold rise in the incidence of mpox was observed in areas of the DRC (Rimoin et al., 2010). In 2017, mpox-associated anogenital lesions were first documented in Nigeria (Ogoina et al., 2019; Yinka-Ogunleye et al., 2019); however, this shift in clinical presentation and transmission was not widely recognized until the global outbreak of mpox Clade IIb caused the declaration of a public health emergency of international concern (PHEIC) in July of 2022 (World Health Organization, 2022). This outbreak was significant because men, and most predominantly those who identified as gay, bisexual and other men who have sex with men (gbMSM), were disproportionately affected (Thornhill et al., 2022). Furthermore, a high prevalence of anogenital lesions was documented, reflecting a documented shift in transmission to intimate contact-based routes (Thornhill et al., 2022). A PHEIC was declared for the second time in August 2024 due to the emergence and subsequent rapid increase of mpox Clade Ib cases in the DRC (World Health Organization, 2024). Unlike 2022, heterosexual transmission, particularly among female sex workers, was of significance during this outbreak (Katoto et al., 2024). Additionally, an outbreak of Clade IIb mpox in Sierra Leone during 2025 resulted in historic infections linked to heterosexual transmission (Mitja et al., 2025), contrasting epidemiological trends of the 2022 global outbreaks. These observations highlight the complex epidemiology of mpox across different geographic locations and population niches.

A contributing factor behind the overall increase in mpox cases may have been the cessation of global smallpox vaccination campaigns. Variola virus (VARV), the causative agent of smallpox, and MPXV are both DNA viruses of the *Orthopoxvirus* genus. *Orthopoxviruses* share conserved genome and amino acid sequences in immunodominant viral proteins, meaning they are serologically cross-reactive (J. Liu et al., 2025). Therefore, residual immunity from Dryvax, the vaccine used during smallpox eradication campaigns, may have prevented the earlier expansion of mpox among human populations (Crandell et al., 2025; Van Dijck et al., 2023). Following the declaration of the eradication of smallpox in 1980, and the subsequent cessation of routine smallpox immunizations, an increasingly immune naïve population has emerged which may be driving increases in mpox cases (Rimoin et al., 2010; Strassburg, 1982).

As of August 2026, Canada has reported 2,381 mpox cases (Public Health Agency of Canada, 2026b) since the beginning of the 2022 outbreaks. The majority of these cases were in Ontario (n = 1,259), Quebec (n = 644), and British Columbia (n = 385) (Public Health Agency of Canada, 2026b). While a smaller number of cases have been reported elsewhere in Canada, the true number of cases is likely higher than reported. The potential stigma arising from identifying as a member of the 2SLGBTQIA+ (Two Spirit, lesbian, gay, bisexual, transgender, queer, intersex, asexual, and other sexually and gender diverse identities) community may have in turn influenced an individual’s willingness to access care (Y. Liu et al., 2025). As opposed to cities such as Toronto and Vancouver, this phenomenon may be more pronounced within conservative provinces, such as the Canadian Prairies, which is composed of Manitoba, Saskatchewan and Alberta. Therefore, while 2 mpox cases were reported in Manitoba (both travel-related), 6 in Saskatchewan, and 71 in Alberta (Public Health Agency of Canada, 2026b), it is possible that the true extent of the outbreak may be greater than recorded.

Within Canada, conservative values may have played a role in the uptake of the mpox vaccine. Canada’s mpox response relied on the Modified Vaccinia Ankara-Bavarian Nordic (MVA-BN) vaccine, a third-generation smallpox vaccine consisting of an attenuated, replication-deficient strain of vaccinia virus (VACV) (Public Health Agency of Canada, 2026a). While initially developed against smallpox, MVA-BN protects against mpox due to extensive serological cross-reactivity among *Orthopoxviruses* (Earl et al., 2004). Earlier smallpox vaccines, such as Dryvax and ACAM2000®, offer this same cross-protection, however, as both rely on live, replication-competent vaccinia strains, they are contraindicated for certain populations and have since been phased out of regular use (Artenstein, 2008; Public Health Agency of Canada, 2026a). As a result, the MVA-BN vaccine became the recommended option, however, owing to limited global supplies, it was initially reserved for only those at high risk of infection (Public Health Agency of Canada, 2022). In response to this, many provinces turned to identity-based vaccine eligibility criteria and dose-sparing strategies, prioritizing men who have sex with other men and sex workers, regardless of gender identity (Frey et al., 2015; Public Health Agency of Canada, 2022). Consequently, this approach required individuals to disclose their gender identity and sexual orientation to a healthcare provider in order to receive the vaccine. However, 2SLGBTQIA+ individuals may delay seeking healthcare or avoid disclosure altogether due to the anticipation of adverse outcomes (Brooks et al., 2018; Seelman et al., 2017). In the Canadian Prairies, conservative values may have suppressed vaccine uptake further.

Assessing the proportion of the population with asymptomatic and/or undetected mpox exposures is crucial to identifying gaps in surveillance and determining the true extent of outbreaks. To date, no mpox serosurveillance in Canada has been published and of those studies conducted among Western countries, sampling has been concentrated within heavily urbanized areas. To investigate evidence of cryptic mpox transmission within key populations residing in conservative regions in Canada and to assess the proportion of the population with *Orthopoxvirus* antibodies, we conducted a cross-sectional seroprevalence study. From June 2024 to December 2025, we collected dried blood spots (DBS) from key populations at community-based organizations and sexual health centers in Manitoba and Alberta, Canada. DBS samples were then analyzed using the Meso Scale Discovery V-PLEX IgG Orthopoxvirus kit to assess seroprevalence using multiple viral antigens.

## Methods

### Ethics statement

Ethical approvals were obtained from both the Committee for Harmonized Health Impact, Privacy, and Ethics Review Health Research Ethics Board (IRB00012696) at the University of Manitoba, and the University of Alberta (Pro00146978), in accordance with university policies. All potential participants were made aware of the study rationale and potential harms prior to providing informed, written consent.

### Study population

Individuals were eligible to participate in this study if they identified as potentially eligible to receive the mpox vaccine, were ≥18 years of age and resided in Manitoba, Saskatchewan or Alberta. This eligibility included, but was not strictly limited to, gbMSM, transgender men and women, non-binary and other gender-diverse individuals, people who have any sexual contact with the individuals described above, as well as sex workers, regardless of their gender. Those with a previously diagnosed mpox infection were excluded from the study.

### Recruitment

Potential participants were recruited via social media, 2SLGBTQIA+ serving community-based organizations (CBOs), sexual health centers and in-person Pride events from June 2024 to December 2025. Additionally, take-home sample collection kits were offered to allow participants to collect their own DBS and mail the sample directly to the laboratory. Alongside their DBS, participants provided brief demographic information (province of residence, age, gender identity, sexual orientation, if they identified as a sex worker, and if they had received any dose of the mpox vaccine).

Recruitment for this seroprevalence study occurred alongside a parallel qualitative study assessing barriers to mpox vaccination among the same population. Qualitative interviews facilitated trust between the research team and marginalized populations, giving an opportunity for research participants to suggest additional sampling avenues that the research team further explored. For instance, in Manitoba, under the guidance of qualitative participants, we recruited at a community health center that provides treatment to people living with HIV and distributes harm reduction supplies to the broader community.

As mentioned above, individuals from any of the Canadian Prairie provinces were eligible to participate, however, despite extensive efforts, no samples were collected from Saskatchewan. At the time of sampling in Saskatchewan, CBOs were actively engaged in advocacy against anti-trans legislation, potentially limiting their ability to participate in research (Walker & Adesanya, 2024).

Additionally, the high burden of communicable diseases (HIV, syphilis, etc.) in Saskatchewan may have hindered physicians’ involvement in this study (Saskatchewan Ministry of Health, 2026). In response to this, sampling efforts were solely focused within Manitoba and Alberta.

### Dried blood spot collection

During in-person participant enrollment events, DBS were collected by trained individuals, or if the individual preferred, they could perform the procedure themselves. If participants chose the latter, detailed instructions on proper sample collection were provided. Capillary blood was collected using a 2 mm contact-activated lancet and dried on Whatman 903 protein saver cards (Cytiva #10534612).

Samples were transported to the laboratory, or arrived via mail, and dried overnight before storage at −20°C. After testing multiple elution and dilution methods, as well as visually assessing the quality of dried blood spots, (**Figure S1**), a ¼ inch hole punch was used to punch out one dried blood spot. This one spot was subsequently eluted in 100 µL of phosphate-buffered saline (PBS; Gibco #10010023) overnight at 4°C. After elution, samples were centrifuged at a minimum of 5,000 x g for 20 minutes. Eluted samples were stored at −80°C. Prior to Meso Scale Discovery (MSD) analysis, eluted samples were further diluted 1:100 in MSD Diluent 100 (MSD, #R50AA-2).

### Orthopoxvirus antigen seroreactivity

Eluted DBS were analyzed using the MSD V-PLEX Orthopoxvirus IgG panel (MSD, #K15688U-4). This multiplexed electrochemiluminescence assay detects antibodies to 5 homologous MPXV and VACV antigens (MPXV A29, MPXV A35, MPXV B6, MPXV E8, MPXV M1 and VACV A27, VACV A33, VACV B5, VACV D8, VACV L1).

Briefly, MSD Blocker A was added to MSD plates for 30 minutes with agitation at 700 RPM and at room temperature (RT). Plates were then washed using MSD Wash Buffer three times (150 µL/well). A four-fold serial dilution was generated using calibrator 1 in Diluent 100. Following this, 50 µL of diluted samples and controls were added to the MSD plates. The plate was then incubated at RT for 2 hours with agitation at 700 RPM, followed by three washes (150 µL/well). Then 50 µL of MSD detection antibody (MSD SULFO-TAG Anti-human IgG Antibody) was added to each well. The plate was subsequently incubated for one hour at RT (with agitation at 700 RPM), followed by three washes (150 µL/well). The MSD GOLD Read Buffer B was then added (150 µL/well) and samples were analyzed using the MESO QuickPlex SQ 120MM. A four-parameter logistic model was then used to calculate the arbitrary units (AU) per mL of each sample from its associated raw signal.

In addition to the provided MSD assay controls, in-house controls were used to validate experimental results. Convalescent serum from PCR+ mpox-infected individuals was used as a positive control for natural infection, while serum and DBS from individuals who were fully vaccinated with MVA-BN was used as a positive control for vaccination. A cohort of seven Canadian individuals with no known *Orthopoxvirus* exposures (no travel to mpox endemic regions and no reported mpox and/or smallpox vaccination or infection) and who were not at high-risk of mpox infection served as negative controls (**Table S1**). This negative cohort was subsequently used to determine seroreactivity cut-offs.

### Cohort stratification

To account for the impact of cross-reactive smallpox vaccination antibodies, serological samples were stratified into four cohorts (Dryvax vaccinated, MVA-BN vaccinated, combination vaccination, and non-vaccinated) based on self-reported MVA-BN vaccination and age. In accordance with the World Health Organization’s declaration of the eradication of smallpox in 1980, participants born prior to 1980 (≥ 45 years of age) were presumed to be Dryvax vaccinated. Those born after 1980 (< 45 years of age) with self-reported mpox vaccination were classified as MVA-BN vaccinated. Participants aged ≥ 45 years of age (indicative of potential Dryvax vaccination) and who reported MVA-BN vaccination were classified as combination vaccinated. Participants born after 1980 and who did not report MVA-BN vaccination were classified as non-vaccinated. Non-vaccinated individuals were analyzed separately from those with *Orthopoxvirus* vaccination as serological cross-reactivity between *Orthopoxviruses* occludes determining if vaccinated individuals have experienced further exposures to mpox.

### Seroreactivity cut-offs

Seroreactivity cut-offs were defined as the mean of negative controls + three standard deviations (**Table S1**). Samples were classified as seroreactive if the concentration of an individual antigen exceeded the corresponding cut-off value. All samples seroreactive to ≥ 6 individual antigen cut-offs were further analyzed for seropositivity. The ratio of orthologous MPXV and VACV antibody concentrations was calculated and samples with a ratio greater than one to a combination of three or more antigen pairs were deemed seropositive. Due to high response variability and a lack of significant differences, the M1R/L1R ratio was excluded from seropositivity assessments. Thus, only ratios of A29L/A27L, A35R/A33R, B6R/B5R and E8L/D8L were included in seropositivity analysis due to previous findings on their utility (Hicks et al., 2024; Pettke et al., 2025; Reed et al., 2026).

### Statistical analysis

Data visualization and statistical analysis was performed using R studio (version 4.3.3). All figures, unless otherwise stated, display calculated concentration as raw values. Geometric mean concentrations (GMC) were further calculated and visualized to assess central tendency. All statistical analyses were performed on log-transformed data. A paired t-test was used to compare differences between orthologous antigen pairs, while a one-way ANOVA with Tukey’s HSD post hoc test compared differences in serological responses induced by potential Dryvax vaccination, MVA-BN vaccination, and combination vaccinationsss. Levels of significance are as follows: *p < 0.05, **p < 0.01, ***p < 0.001, ****p < 0.0001. Additional R packages used during data manipulation and analysis include tidyr v1.3.1 (Wickham et al., 2024), dplyr v1.1.4 (Wickham et al., 2023), ggplot2 v4.0.0 (Wickham, 2016), ggbeeswarm v0.7.3 (Clarke et al., 2025), ggpubr v0.6.3 (Kassambara, 2026), rstatix v0.7.3 (Kassambara, 2025), UpSetR v1.4.0 (Gehlenborg, 2019), patchwork v1.3.2 (Pedersen, 2025) and gtsummary v2.4.0 (Sjoberg et al., 2021).

## Results

### Participant Demographics

From June 2024 to December 2025, 275 individuals were recruited for this study (**Table 1**). The median age of participants was 31 years (IQR = 25, 40), with ages ranging from 18-75 years. Most individuals identified as gay (31.3%, n = 86) and as men (56.7%, n = 159). Respondents were similarly distributed between Alberta (49.8%; n = 137) and Manitoba (50.2%; n = 138). In Alberta, those who identified as gay (45.3%, n = 62) and as men (70.1%, n = 96) made up the largest proportion of participants. In Manitoba, men represented 43.5% (n = 60) of provincial participants, while women represented 34.1% (n = 47). There were slight differences regarding sexual orientation in Manitoba, with bisexual (29.0%, n = 40) and heterosexual (29.7%, n = 41) identifying individuals comprising most of the respondents. These differences may partially be explained by the higher number of sex workers recruited in Manitoba. Forty-five individuals who identified as sex workers were recruited overall, with 91.1% residing in Manitoba (41/45) and 8.9% residing in Alberta (4/45). Of those who identified as sex workers, 40.0% (n = 18) identified as bisexual and 26.7% (n = 12) identified as heterosexual. Further differences among participant demographics may have been influenced by sampling strategies between provinces.

**Table 1.**
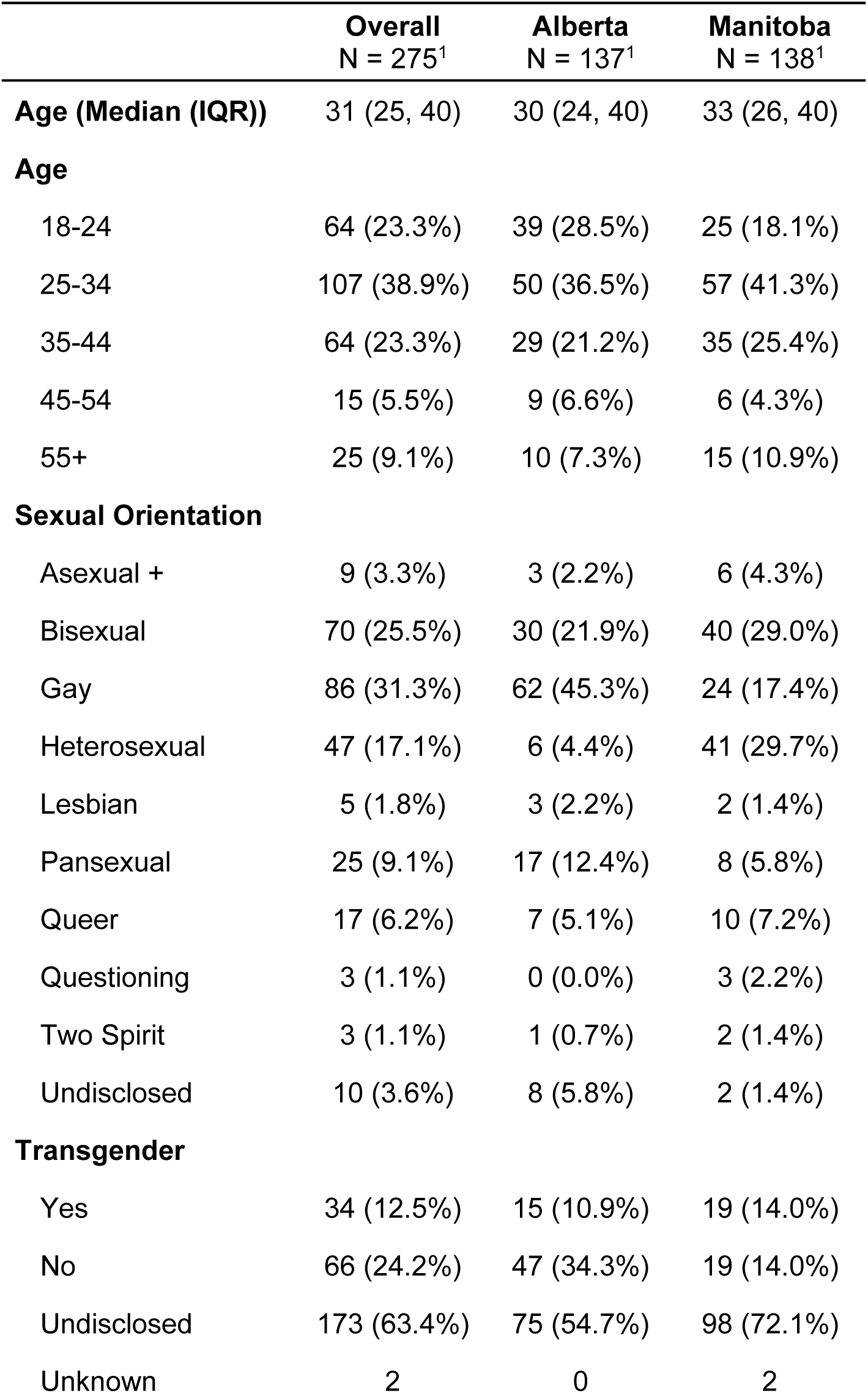
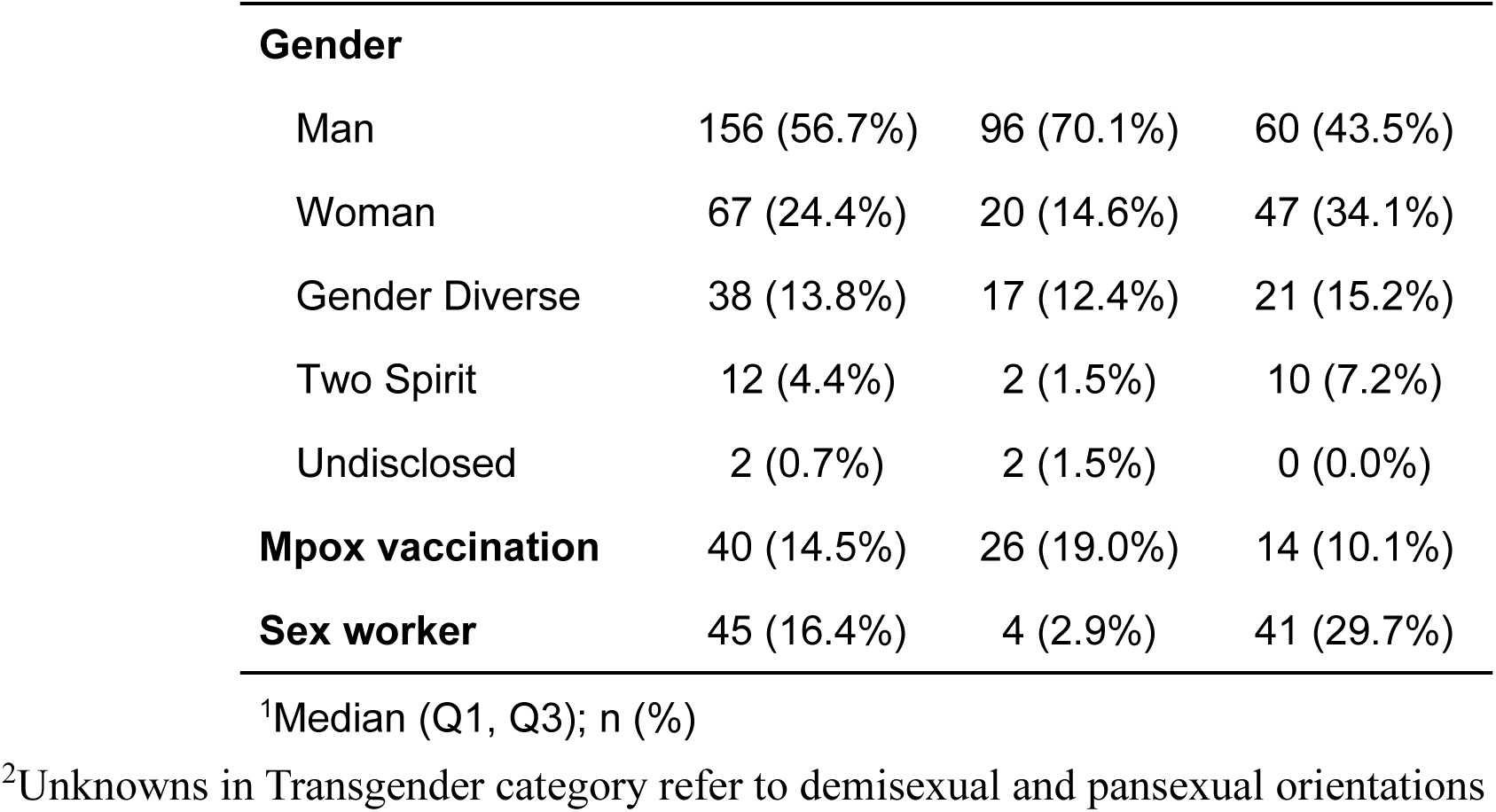
Demographics of all participants.

### Mpox vaccination rates

Of all participants recruited, 14.5% (n = 40) self-reported receiving at least one dose of the mpox vaccine (**Table 2**). Most vaccinated individuals were cisgender (41.0%, n = 16), gay (52.5%, n = 21) and identified as a man (65.0%, n = 26). The median age of vaccinated individuals was 36 years (IQR = 30, 43). Stark differences arose when comparing the number of vaccinated individuals between provinces, with 65.0% (n = 26) of vaccinated individuals residing in Alberta and 35.0% (n = 14) residing in Manitoba. As previously mentioned, differences in sampling may have also influenced this difference. Among the 45 individuals who identified as sex workers, only two individuals reported receiving any dose of the mpox vaccine (4.4%).

**Table 2.** Characteristics of unvaccinated and MVA-BN vaccinated individuals.

|  | **Unvaccinated**<br>N = 235<sup>1</sup> | **Vaccinated**<br>N = 40<sup>1</sup> |
| --- | --- | --- |
| **Province** |  |  |
| Alberta | 111 (47.2%) | 26 (65.0%) |
| Manitoba | 124 (52.8%) | 14 (35.0%) |
| **Age (Median (IQR))** | 30 (24, 39) | 36 (30, 43) |
| **Age** |  |  |
| 18-24 | 59 (25.1%) | 5 (12.5%) |
| 25-34 | 93 (39.6%) | 14 (35.0%) |
| 35-44 | 51 (21.7%) | 13 (32.5%) |
| 45-54 | 11 (4.7%) | 4 (10.0%) |
| 55+ | 21 (8.9%) | 4 (10.0%) |
| **Sexual Orientation** |  |  |
| Asexual + | 8 (3.4%) | 1 (2.5%) |
| Bisexual | 65 (27.7%) | 5 (12.5%) |
| Gay | 65 (27.7%) | 21 (52.5%) |
| Heterosexual | 44 (18.7%) | 3 (7.5%) |
| Lesbian | 5 (2.1%) | 0 (0.0%) |
| Pansexual | 24 (10.2%) | 1 (2.5%) |
| Queer | 12 (5.1%) | 5 (12.5%) |
| Questioning | 3 (1.3%) | 0 (0.0%) |
| Two Spirit | 2 (0.9%) | 1 (2.5%) |
| Undisclosed | 7 (3.0%) | 3 (7.5%) |
| **Transgender** |  |  |
| Yes | 28 (12.0%) | 6 (15.4%) |
| No | 50 (21.4%) | 16 (41.0%) |

|  | Unvaccinated<br>N = 235<sup>1</sup> | Vaccinated<br>N = 40<sup>1</sup> |
| --- | --- | --- |
| Undisclosed | 156 (66.7%) | 17 (43.6%) |
| Unknown | 1 | 1 |
| **Gender** |  |  |
| Man | 130 (55.3%) | 26 (65.0%) |
| Woman | 61 (26.0%) | 6 (15.0%) |
| Gender Diverse | 35 (14.9%) | 3 (7.5%) |
| Two Spirit | 8 (3.4%) | 4 (10.0%) |
| Undisclosed | 1 (0.4%) | 1 (2.5%) |
| **Sex worker** | 43 (18.3%) | 2 (5.0%) |

<sup>1</sup>n (%); Median (Q1, Q3)

### Orthopoxvirus serological responses

A total of 274 DBS samples were analyzed via MSD. Significant differences in the GMC across all participants were found between MPXV A29 and VACV A27 (21.11 vs. 19.23, p < 0.0001), MPXV A35 and VACV A33 (41.45 vs. 36.29, p < 0.0001), as well as MXPV B6 and VACV B5 (30.60 vs. 32.12, p < 0.0001) (**Figure 1**; **Table 3**). There were no statistically significant differences between the GMC of MPXV E8 and VACV D8, in addition to MPXV M1 and VACV L1.

**Figure 1.**
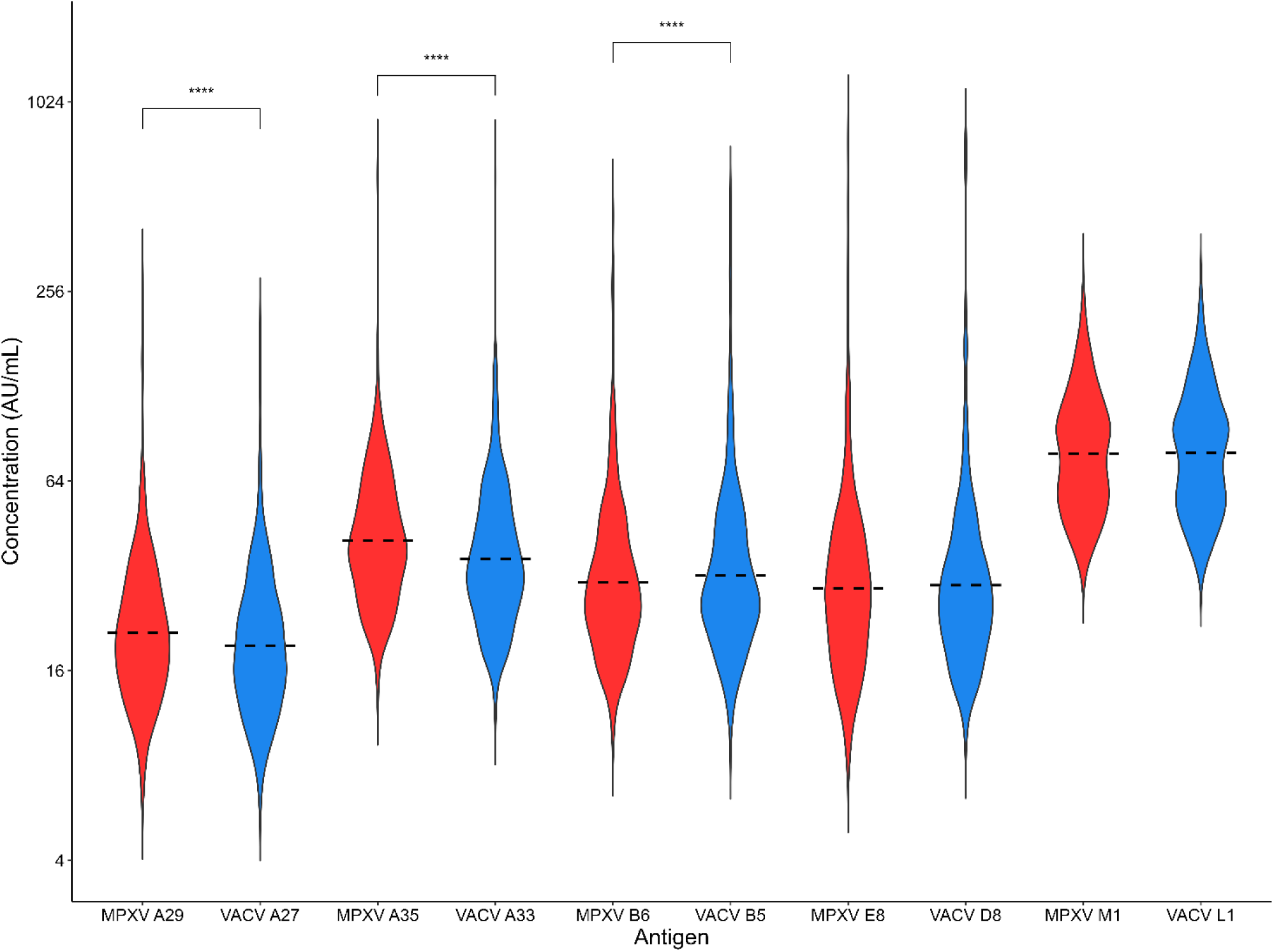
Serological responses of homologous MPXV and VACV antigens within the whole sample (n = 274). MPXV and VACV antigens are shown in red and blue, respectively. Antibody concentrations depicted as untrimmed violin plots with overlaid geometric means (dashed horizontal lines). Statistical differences assessed via paired t-test using log-transformed data (**** p < 0.0001). Only statistically significant differences shown.

**Figure 2.**
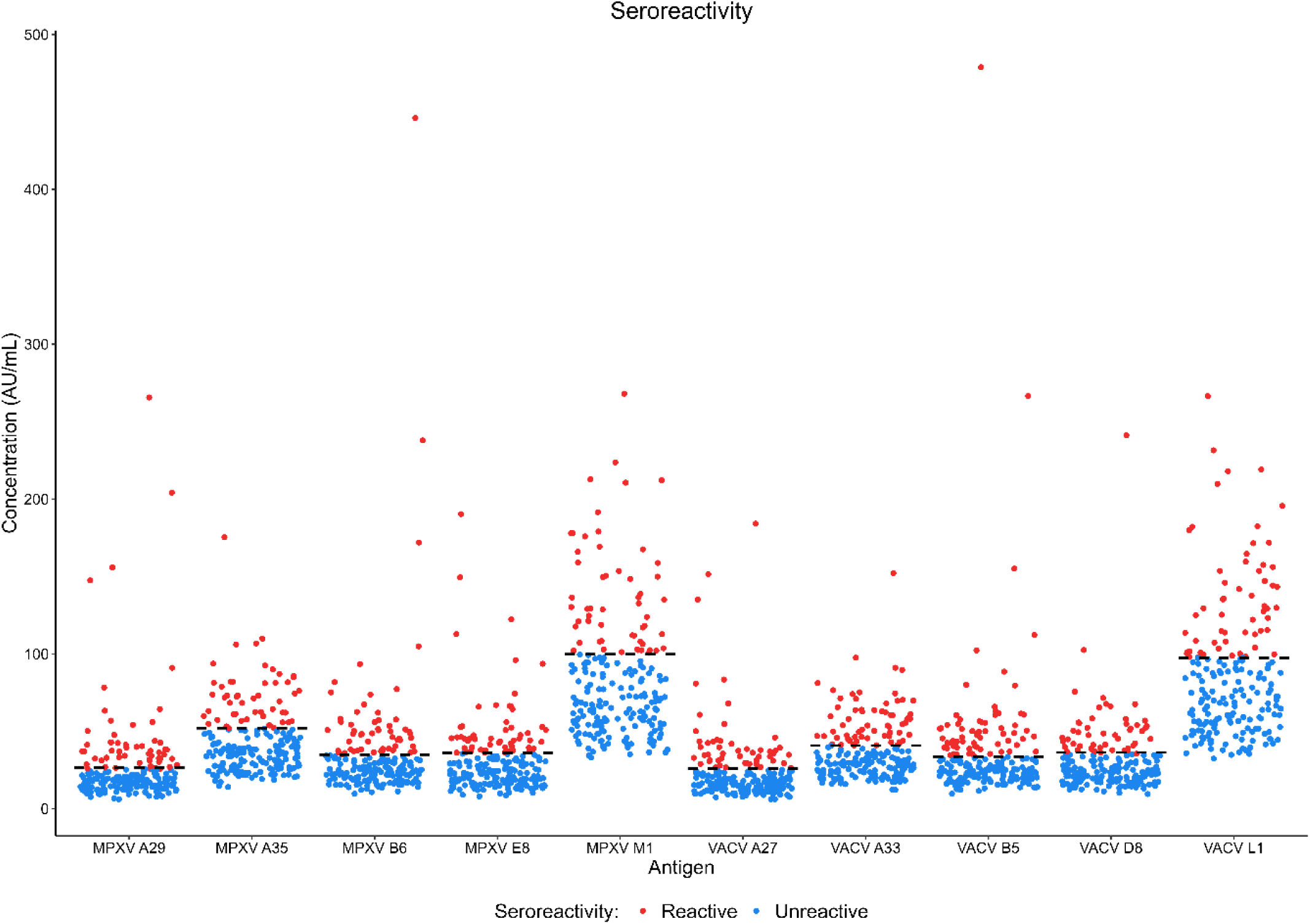
**Classification of samples from individuals with no prior *Orthopoxvirus*** vaccination as seroreactive using cut-off values (n = 203). Individual antigen cut-offs are displayed as horizontal dashed lines. Seroreactive samples, those with concentrations higher than the cut-off, are depicted as red while those lower than the cut-offs are depicted as blue.

**Table 3.** Geometric mean antibody concentrations (GMC) of homologous MPXV and VACV antigens across the whole sample, as well as geometric standard deviation (GSD). Differences between homologous antigens assessed via paired t-test.

| *Homologous antigen*<br>*pairs* | *GMC (95% CI)* | *GSD* | *p-value* |
| --- | --- | --- | --- |
| *MPXV A29*<br>*VACV A27* | 21.08 (19.79-22.45)<br>19.19 (18.04-20.41) | 1.70<br>1.68 | p < 0.0001 |
| *MPXV A35*<br>*VACV A33* | 41.39 (39.15-43.77)<br>36.25 (34.14-38.48) | 1.60<br>1.65 | p < 0.0001 |
| *MPXV B6*<br>*VACV B5* | 30.57 (28.59-32.69)<br>32.09 (30.00-34.36) | 1.76<br>1.77 | p < 0.0001 |
| *MPXV E8*<br>*VACV D8* | 29.23 (27.08-31.55)<br>29.88 (27.72-32.21) | 1.90<br>1.88 | p = 0.4 |
| *MPXV M1*<br>*VACV L1* | 78.32 (74.43-82.40)<br>78.52 (74.58-82.67) | 1.53<br>1.54 | p = 0.46 |

### Seroreactivity among non-vaccinated individuals

After stratification by vaccination status and age, 203 individuals (73.8%) were classified as non-vaccinated. In total, 21.7% (n = 44) were considered seroreactive to ≥ 6 individual antigenic cut-offs. The majority of these seroreactive samples (15.8%; n = 32) had a concentration greater than the cut-off value for all ten antigens present (**Figure S2**). The 44 seroreactive samples were then further classified as seropositive using the ratio of MPXV to VACV antibodies. All these seroreactive samples had a ratio > 1 for A35/A33 (**Figure 3**). Following this, there were 34 samples with a ratio >1 for A29/A27, 12 with a ratio > 1 for B6/B5 and only 9 samples with a ratio greater >1 for E8/D8 (**Figure 3**).

**Figure 3.**
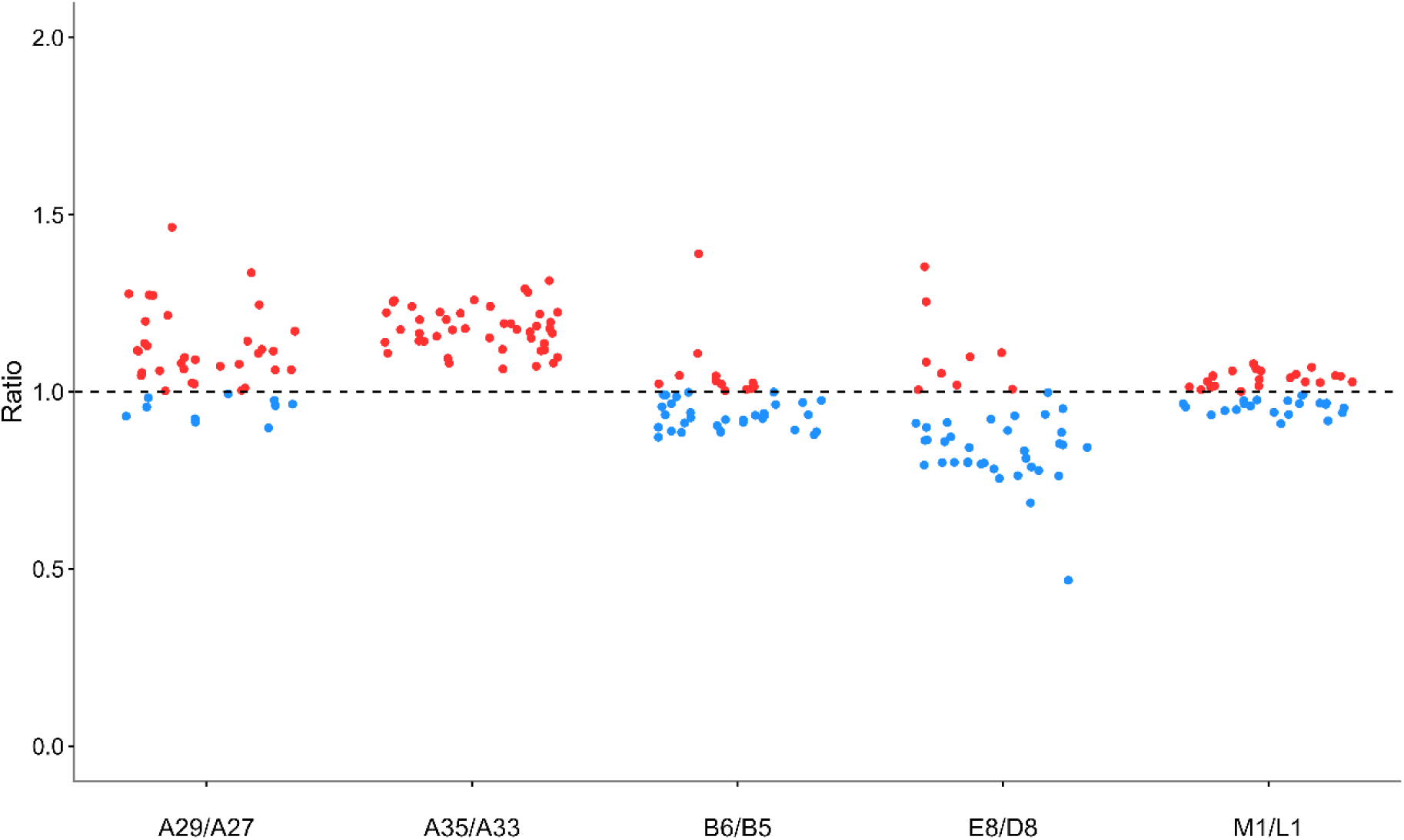
MPXV to VACV antigen concentration ratios from seroreactive samples (n = 44). Samples with a ratio >1 are depicted as red and those with a ratio <1 are depicted as blue.

When assessing seropositivity through combinations of homologous antigens, only 2 samples were seropositive using all four antigen pairs (A29/A27, A35/A33, B6/B5 and E8/D8). An additional ten samples were seropositive using any combination of three antigen pairs (**Figure 4**). Therefore, in total, 5.9% (12/203) of the samples were seropositive to MPXV antigens (**Figure 5**).

**Figure 4.**
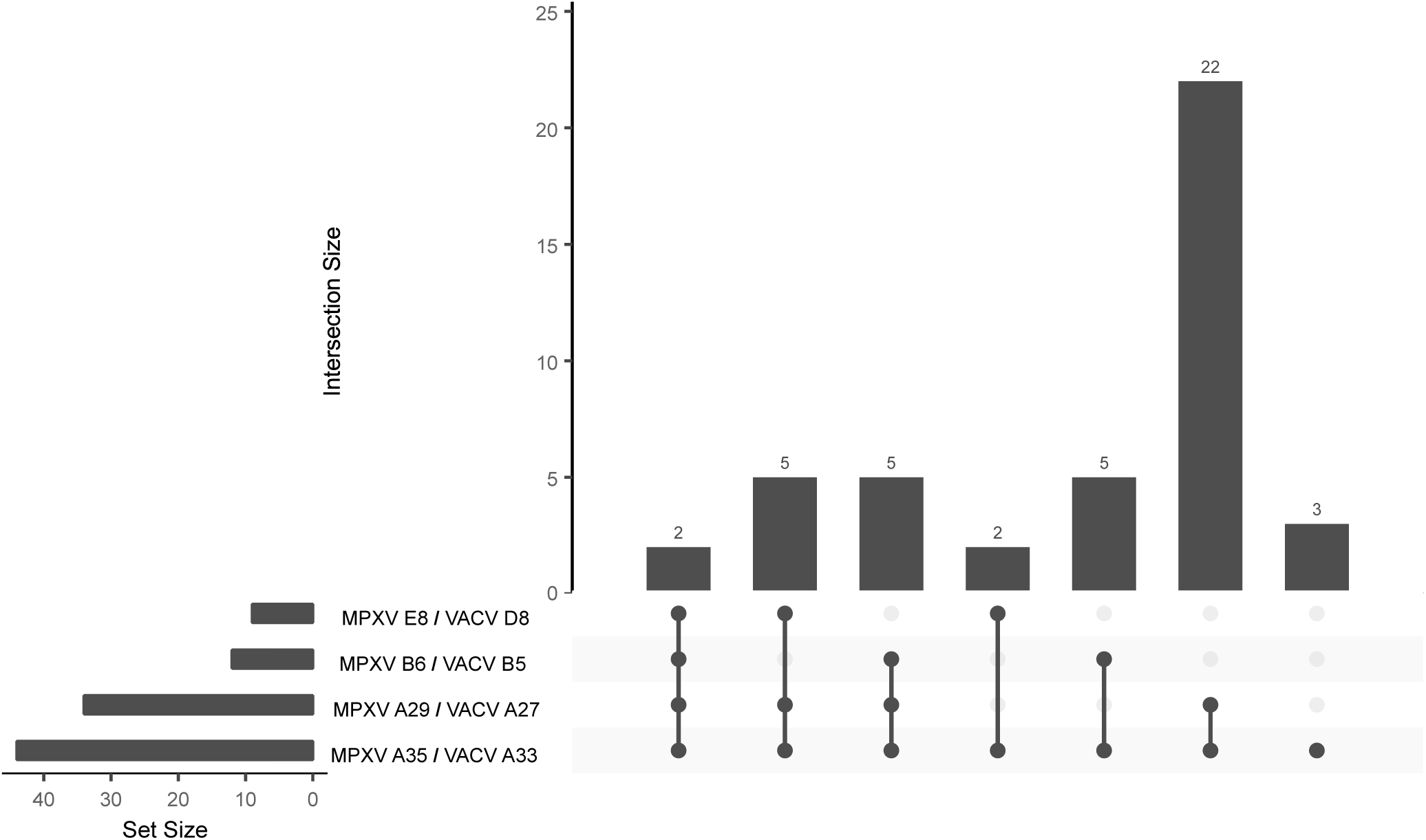
Upset plot of MPXV seropositivity. Horizontal bar graphs display absolute number of samples with a ratio >1 for each individual antigen pair. Sample seropositivity (as determined by the ratio of MPXV to VACV) is depicted as combinations of antigen pairs through vertical bar graphs.

**Figure 4.**
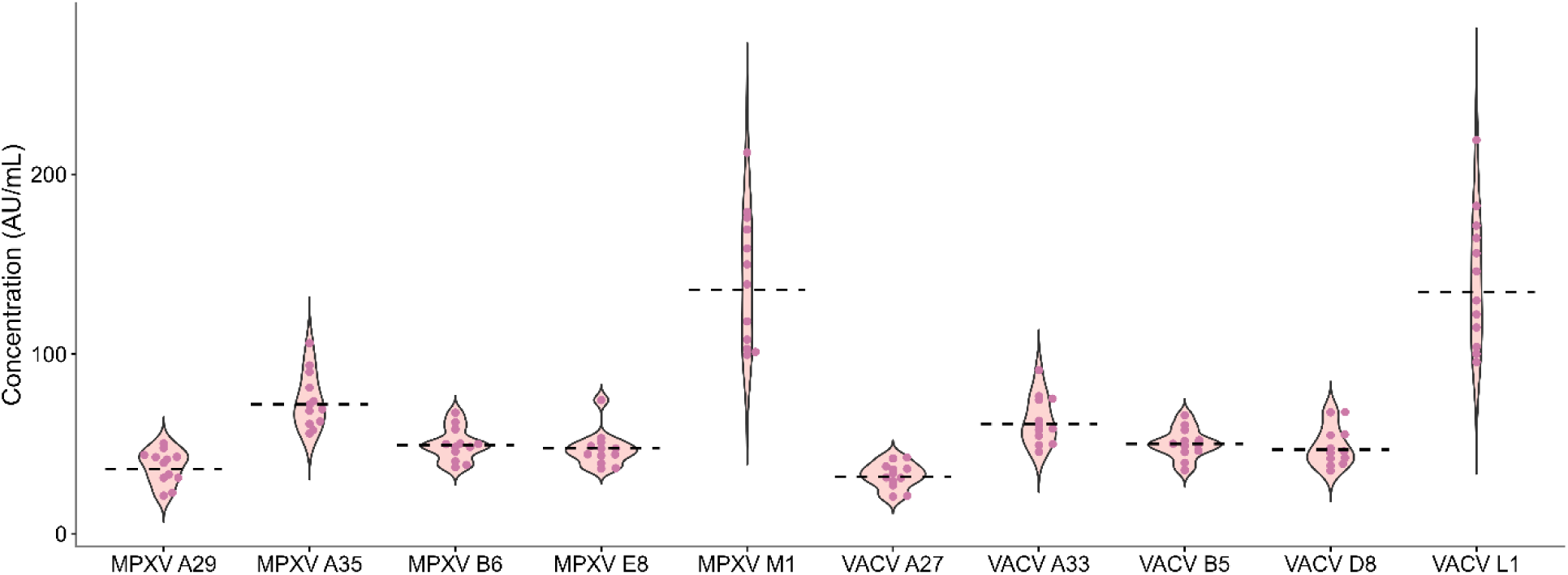
Seropositive samples as determined through antigenic cut-offs and subsequent MPXV:VACV ratios (n = 12). Geometric mean concentrations are displayed for each antigen as dashed horizontal lines.

**Figure 5.**
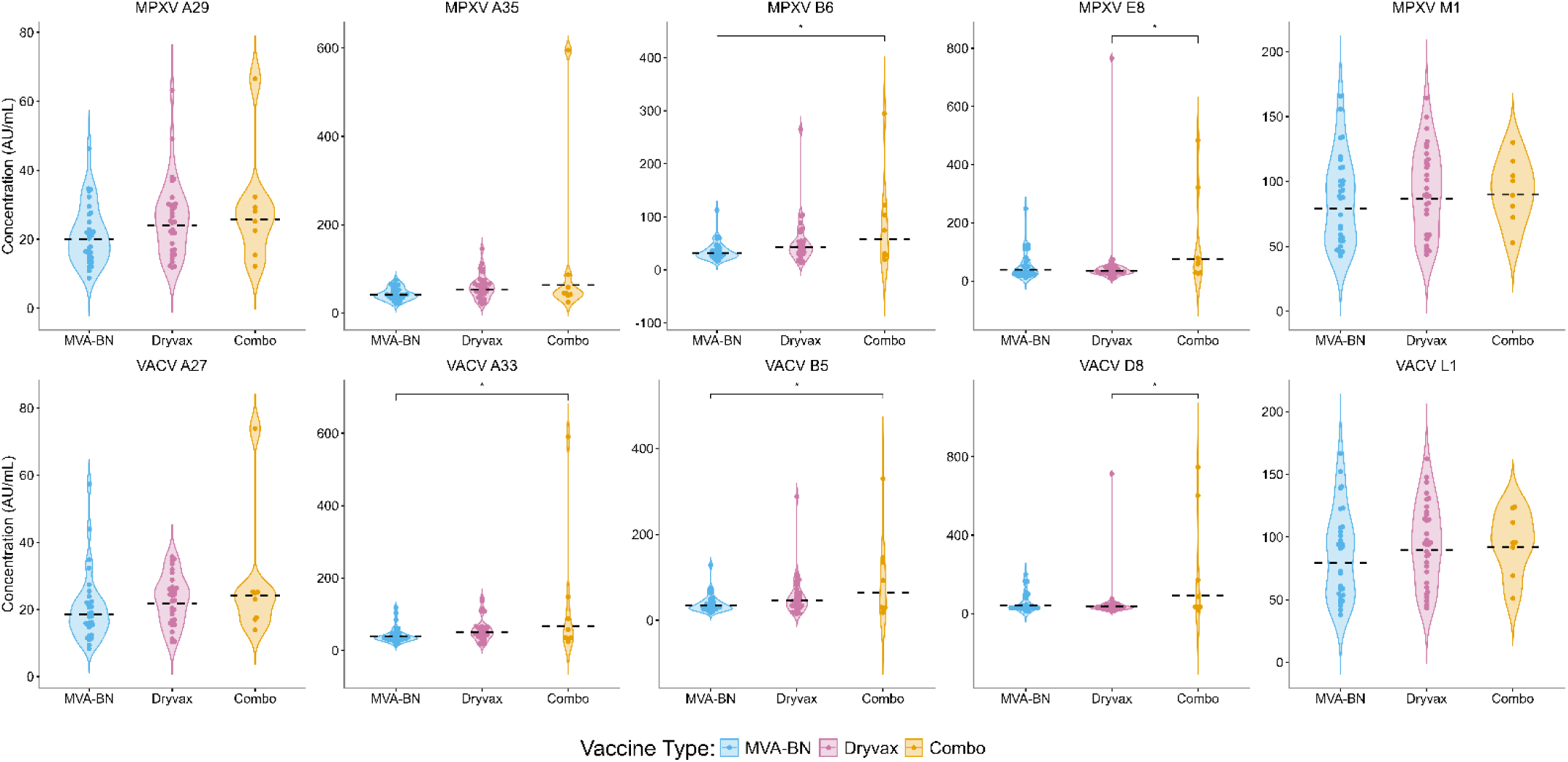
Serological responses among individuals with *Orthopoxvirus* vaccination. Individuals were classified based on age and MVA-BN vaccination status. Individuals who reported MVA-BN vaccination and who were born post 1980 (n=31) are shown in blue, individuals who were born prior to 1980 (potential Dryvax vaccination) (n=32) are shown in pink, while a small subset of individuals (n=8) born prior to 1980 who also reported receiving the MVA-BN vaccine are shown in yellow (Combo vaccination). The calculated concentration of antibody levels (AU/mL) are shown on the y-axis and GMCs are represented as dashed horizontal lines. Original concentration values were log-transformed for statistical analysis. Only significant results (as determined by ANOVA and Tukey’s HSD post hoc) are shown (* p < 0.05).

### Misalignment between seropositive individuals and vaccine eligibility

Of the twelve individuals who were classified as seropositive, a multitude of gender identities and sexual orientations were reported. Regarding gender, cisgender and transgender men and women, as well as Two Spirit, and gender diverse individuals were seropositive. The sexual orientations of those who were seropositive included heterosexual, bisexual, gay, and queer. Thus, while most vaccinated individuals identified as cisgender gay men, a broader range of identities were captured among seropositive individuals. Geographically, four individuals were from Alberta, while nine resided in Manitoba. Age distribution was also varied, four individuals were between the ages of 18-24, three were aged 25-34 years and five were aged 35-44 years. Lastly, a small number of individuals who were seropositive identified as sex workers (2/12; 4.45% of sex workers overall). Between provinces, most vaccinated individuals resided in Alberta; however, more seropositive individuals were located in Manitoba.

### Serological responses among vaccinated individuals

To compare serological differences induced by first and third-generation *Orthopoxvirus* vaccines, individuals were classified into three cohorts based on self-reported MVA-BN vaccination and age. There were 31 individuals aged < 45 years with MVA-BN vaccination, 32 individuals aged ≥ 45 years with potential Dryvax vaccination and 8 individuals ≥ 45 years with both MVA-BN and potential Dryvax vaccination.

Consistently, a trend was observed wherein the highest GMCs were recorded amongst those who received both Dryvax and MVA-BN vaccines, followed by those with only Dryvax vaccination (**Figure 6**; **Table 4**). However, among those who have received the MVA-BN vaccine, elevated levels of MPXV E8 and VACV D8 are observed in comparison to those who have received the Dryvax vaccine (E8 = 39.39 vs. 35.90 p = 0.88; D8 = 43.46 vs. 36.26, p = 0.65). Despite these trends, statistically significant differences between cohorts were restricted to A35/A33, B6/B5 and E8/D8. Specifically, individuals with both MVA-BN and Dryvax vaccination had higher antibody levels than those with only MVA-BN vaccination for MPXV B6 (57.70 vs. 32.35, p < 0.05), and compared with those with only Dryvax vaccination for MPXV E8 (75.05 vs. 35.90, p < 0.05). Similar patterns were observed when assessing serological responses to VACV. When assessing antibodies to VACV B5, individuals with a combination of vaccines demonstrated higher levels of antibodies in comparison to those with only MVA-BN vaccination (63.72 vs. 35.08, p < 0.05). Similarly, those with a combination of vaccines had higher levels of antibodies to VACV D8 in comparison to those with only Dryvax vaccination (90.65 vs. 36.26, p < 0.05). Unlike its orthologous pair, those who had received both Dryvax and MVA-BN vaccines had higher antibody levels than those with only MVA-BN vaccination for VACV A33 (67.50 vs. 38.27, p < 0.05).

**Table 4.** Antibody responses of various *Orthopoxvirus* based vaccines. Individuals < 45 years of age and self-reported mpox vaccination were classified as MVA-BN vaccinated (n =31). Those ≥ 45 years of age with no self-reported mpox vaccination were classified as Dryvax vaccinated (n=32). Individuals who were ≥ 45 years of age who self-reported mpox vaccination were classified as combination vaccinated (both MVA-BN and Dryvax vaccinated) (n=8). Antibody concentrations are represented via GMC with 95% confidence intervals (95% CI). The GSD is also calculated. Differences in antibody levels between groups were assessed via a one-way ANOVA.

|  | *Antigen* | *MVA-BN vaccination*<br>*(n = 31)* |  | *Dryvax vaccination*<br>*(n=32)* |  | *Combination*<br>*vaccination (n=8)* |  | *Global p-*<br>*value* |
| --- | --- | --- | --- | --- | --- | --- | --- | --- |
|  |  | *GMC*<br>*(95% CI)* | *GSD* | *GMC*<br>*(95% CI)* | *GSD* | *GMC*<br>*(95% CI)* | *GSD* |  |
| *MPXV* | *A29* | 19.89<br>(17.24-22.95) | 1.48 | 23.91<br>(20.63-27.71) | 1.51 | 25.65<br>(16.74-39.31) | 1.67 | p = 0.13 |
|  | *A35* | 40.90<br>(36.32-46.06) | 1.38 | 53.69<br>(45.35-63.56) | 1.60 | 62.88<br>(27.84-142.01) | 2.65 | p = 0.03 |
|  | *B6* | 32.35<br>(27.76-37.69) | 1.52 | 42.99<br>(34.45-53.64) | 1.85 | 57.70<br>(26.05-127-82) | 2.59 | p = 0.027 |
|  | *E8* | 39.39<br>(30.16-51.45) | 2.07 | 35.90<br>(27.69-46.54) | 2.06 | 75.05<br>(29.65-190) | 3.04 | p = 0.059 |
|  | *M1* | 79.23<br>(68.39-91.79) | 1.49 | 86.88<br>(75.63-99.80) | 1.47 | 90.14<br>(70.90-114.61) | 1.33 | p = 0.54 |
| *VACV* | *A27* | 18.54<br>(15.83-21.72) | 1.54 | 21.73<br>(19.16-24.64) | 1.42 | 24.08<br>(15.80-36.72) | 1.66 | p = 0.15 |
|  | *A33* | 38.27<br>(32.66-44.84) | 1.54 | 49.56<br>(40.76-60.27) | 1.72 | 67.50<br>(27.86-163.50) | 3.88 | p = 0.032 |
|  | *B5* | 35.08<br>(29.97-41.07) | 1.54 | 45.81<br>(36.56-57.41) | 1.87 | 63.72<br>(26.86-151.17) | 2.81 | p = 0.034 |
|  | *D8* | 43.46<br>(33.07-57.10) | 2.11 | 36.26<br>(28.51-46.11) | 1.95 | 90.65<br>(27.68-296.91) | 4.13 | p = 0.021 |
|  | *L1* | 79.60<br>(68.36-92.70) | 1.51 | 89.36<br>(78.20-102.11) | 1.45 | 91.79<br>(71.24-118.27) | 1.35 | p = 0.42 |

## Discussion

Using a community-based investigational approach, twelve *Orthopoxvirus* seropositive individuals were identified in our study, suggesting the possibility of undetected mpox transmission within the Canadian Prairies. Furthermore, 44 individuals had antibodies to six or more antigens assessed, further supporting evidence of previously undetected *Orthopoxvirus* circulation. Between provinces, more seropositive individuals resided in Manitoba than Alberta. This contrasts with confirmed mpox cases reported by the Public Health Agency of Canada, with Alberta reporting more cases than Manitoba (Public Health Agency of Canada, 2026b). As discussed by Boutzoukas et al, stigma, whether internal or external, may have influenced health-seeking behaviours among those with prior mpox infection or those at risk of infection in the United States (Boutzoukas et al., 2025), which similarly may have led to an underreporting of mpox cases in Canada. The influence of stigma on health-seeking behaviours was likely exacerbated if individuals resided in politically conservative regions, such as Manitoba and Alberta, which were governed by politically conservative parties at the time of the 2022 mpox outbreaks.

In New York City, a similar rate of seroprevalence (6.4%) was observed among non-vaccinated individuals presenting for care at sexual health clinics (Pathela et al., 2024). Furthermore, 7.37% of non-vaccinated people living with HIV in Rome, Italy who were attending a hospital outpatient service had antibodies against mpox (Salvo et al., 2024). Despite these geographical regions reporting a higher number of mpox cases, similar rates of seroprevalence were observed. Out of the 45 sex workers in our study, two were seropositive (4.45%). A similar rate was observed among sex workers in Thailand (4.6%; 12/262), however it is important to note that sample sizes differed (Umer et al., 2025). Like others, our results also indicate mpox serological exposures were not solely among men who have sex with men (Pathela et al., 2024; Salvo et al., 2024). The broad range of identities, including heterosexual cisgender individuals, suggests more individuals were at risk for mpox infection than those prioritized during vaccine campaigns. Based on their findings, Waddell et al., have also posited that persons experiencing homelessness, regardless of gender or sexual orientation, are also at risk of mpox infection and require equitable access to the mpox vaccine (Waddell et al., 2023). Ultimately, these findings underscore that mpox transmission is driven by specific risk factors rather than identity.

Based on the questions asked during this study, it is unknown how seropositive individuals were exposed to mpox or other *Orthopoxviruses*. Within Canada, there are no known endemic *Orthopoxviruses*, yet surveillance has thus far been very limited. Globally, cowpox and bovine vaccinia are endemic to Europe and Brazil, respectively, with *Orthopoxvirus* antibodies detected among both Finnish veterinarians and Brazilian agricultural workers (Baxby et al., 1994; Chantrey et al., 1999; Costa et al., 2016; Jose da Silva Domingos et al., 2021; Ninove et al., 2009; Pelkonen et al., 2003). In North America, seven cases of borealpox have been identified among people in Alaska, and evidence of *Orthopoxvirus* antibodies have been found in wild small mammals within the same region (Mooring et al., 2026; Springer et al., 2017). These studies highlight how various *Orthopoxviruses*, besides mpox, continue to circulate globally and have the potential to be distributed across Canada. While the absence of adequate *Orthopoxvirus* surveillance in Canada precludes knowing if virus circulation is indeed occurring, having an understanding of the typical geographical range in which cases of *Orthopoxviruses* occur provides additional support for potential mpox exposures in the Canadian Prairies.

Out of all the participants, only 14.5% reported receiving the mpox vaccine. This mirrors low mpox vaccination rates globally (Sulaiman et al., 2024). In the United States, full two-dose mpox vaccine coverage was estimated to be 22% among those with an increased risk of infection (Owens et al., 2023). While the District of Columbia, Washington has been able to reach coverage estimates of 66.2%, traditionally conservative states, such as West Virginia have not been as successful (4.6% coverage) (Owens et al., 2023). In Canada, among 2LSGBTQ+ individuals who were aware of the vaccine in 2023, only 17.3% had received at least one dose (Public Health Agency of Canada, 2026c). While no aggregated provincial level data exists in Canada, we can assess trends in mpox vaccine coverage across specific regions of Canada. In 2023, 12.9% of 2SLGBTQ+ survey respondents in British Columbia and the Canadian Prairies (Manitoba, Saskatchewan, and Alberta) had received at least one dose of the mpox vaccine, in comparison to 18.4% of respondents in Ontario and 21.2% of respondents in Quebec and the Atlantic provinces (Public Health Agency of Canada, 2026c). While this finding demonstrates that the highest mpox vaccine coverage was in Quebec and the Atlantic provinces, it also underscores the importance of provincial level data in Canada to inform local vaccination campaigns.

Low uptake of the mpox vaccine in Canada may have been impacted by complex, oftentimes unclear, identity-based eligibility criteria that necessitated individuals disclosing their gender identity and sexual orientation to their healthcare provider, however, anticipated fear or discrimination may prevent some individuals from doing so, in turn preventing them from receiving the mpox vaccine (Birch et al., 2024; Brooks et al., 2018). Furthermore, the fluidity of sexual orientation and gender identity may also have impacted how individuals discerned if they were eligible for the vaccine. Importantly, vaccine criteria structured around gender and sexual orientation requirements may not entirely encompass those who are at risk of mpox infection as demonstrated by our results. While vaccination campaigns appeared to reach cisgender gay men, of whom the majority resided in Alberta, serological exposures were disproportionately located in Manitoba and not confined to this demographic. Universal mpox vaccine eligibility, in relation to gender and sexual orientation, may potentially better encompass those at risk of mpox infection. Positioning mpox as a concern for only the 2SLGBTQIA+ community within the media and public health messaging increases stigmatization and has the potential to perpetuate cryptic virus transmission within unprioritized populations engaging in high-risk behaviours (Boutzoukas et al., 2025).

In comparison to those with presumed historical smallpox vaccination and recent mpox vaccination, those who had only one type of vaccine (Dryvax or MVA-BN), had lower antibody concentrations across multiple antigens. While only one facet of the immune response was assessed, this supports what others have identified, in that MVA-BN vaccination acts as an immunological booster among those with previous replication-competent vaccination (W. D. Liu et al., 2025; Van Dijck et al., 2026). Furthermore, those with presumed historical Dryvax vaccination demonstrated higher antibody responses than those who have only recently received the MVA-BN vaccine in our study, except against MPXV E8 and VACV D8. This further supports evidence that those with historical Dryvax vaccination still retain circulating antibodies decades after immunization (Taub et al., 2008). This is in direct contrast to the waning antibody response observed post MVA-BN vaccination (Prevost et al., 2026). Our findings also align with recent results that suggest the immune response induced by MVA-BN vaccination may not be as robust or long-lasting as that induced by Dryvax immunization (Crandell et al., 2026). Altogether, the waning humoral responses observed within our study, in conjunction with low vaccination rates, highlights an increasingly immune naïve population at risk of MPXV infection in Canada.

## Limitations

While dual methods of determining seropositivity have been utilized, differentiating *Orthopoxvirus* serological exposures is nuanced and requires population-specific context that is currently lacking in Canada. Therefore, while we have used a group of unexposed Canadians (n = 7) as negative controls, a larger, more representative sample of both known negative and positive controls would strengthen the determination of cut-off values and allow for sensitivity and specificity to be assessed via receiver operating characteristic curves. Furthermore, the cross-reactivity of orthologous antigens and utilization of ratios must be further validated within the target population and setting. For instance, while we first assessed seroreactivity by using the mean + three standard deviation approach, a value greater than one may be required when interpreting the ratio of orthologous antigens, particularly for A35/A33.

Secondly, the role that T-cells play in MVA-BN vaccine-induced immunity is increasingly being demonstrated (Cohn et al., 2023; Grifoni et al., 2022). Consequently, it may be that individuals with low humoral immunity post MVA-BN vaccination in our study may have instead been able to mount a robust cell-mediated immune response. Despite this limitation, dried blood spots offer a logistical advantage over traditional phlebotomy, particularly in a community-based context. Additionally, no virus neutralization assays were performed in this study, and it is unknown how the low humoral immune response demonstrated would translate to a neutralizing antibody response.

Thirdly, the lack of supporting epidemiological data in this study limits what types of conclusions can be ascertained, particularly surrounding routes of *Orthopoxvirus* exposure and risk factors. Furthermore, vaccination rates are subject to desirability and recall biases. In the future, supporting documentation of mpox or smallpox vaccination would strengthen both epidemiological and serological findings. Lastly, the small sample size and restricted geographical area of the study may limit the generalizability of these results.

## Conclusions

Overall, this study successfully demonstrates how community-based seroprevalence studies can provide insights into viral transmission that routine passive surveillance cannot adequately capture. Using this method, we identified twelve potentially undetected *Orthopoxvirus* exposures in Manitoba and Alberta among individuals who did not always conform with restrictive provincial mpox vaccine eligibility. Furthermore, while a durable humoral immune response is maintained among those with presumed historical smallpox vaccination, waning antibody responses were observed among those with MVA-BN vaccination. When this finding is situated alongside the low vaccination rates reported by participants, an increasingly immune naïve population vulnerable to future mpox outbreaks in the Canadian Prairies is highlighted.

Shifting away from identity-based criteria to universal eligibility will minimize stigma, lower barriers to accessing the mpox vaccine, and better protect hidden at-risk populations. Additionally, augmenting traditional vaccination campaigns with community-based approaches may improve mpox vaccine uptake. While increasing vaccine coverage among key populations in Canada is a priority, an even greater gap in vaccine coverage exists in mpox endemic regions of Africa. Unless addressed and remedied, persistent mpox outbreaks will occur with the potential for global expansion. In conclusion, shifting to universal mpox vaccine eligibility, with respect to gender identity and sexual orientation requirements, will eliminate barriers, curb viral transmission, and promote health equity across the Canadian Prairies.

## Acknowledgements

The authors would like to thank all community participants and organizations for their participation in this investigation. This work was funded by the International Mpox Research Consortium (IMReC) through funding from the Canadian Institutes of Health Research and International Development Research Centre (grant no. MRR-184813).

## Data Availability

Data is available upon reasonable request to the authors.

## Supplemental Tables and Figures

**Figure S1.**
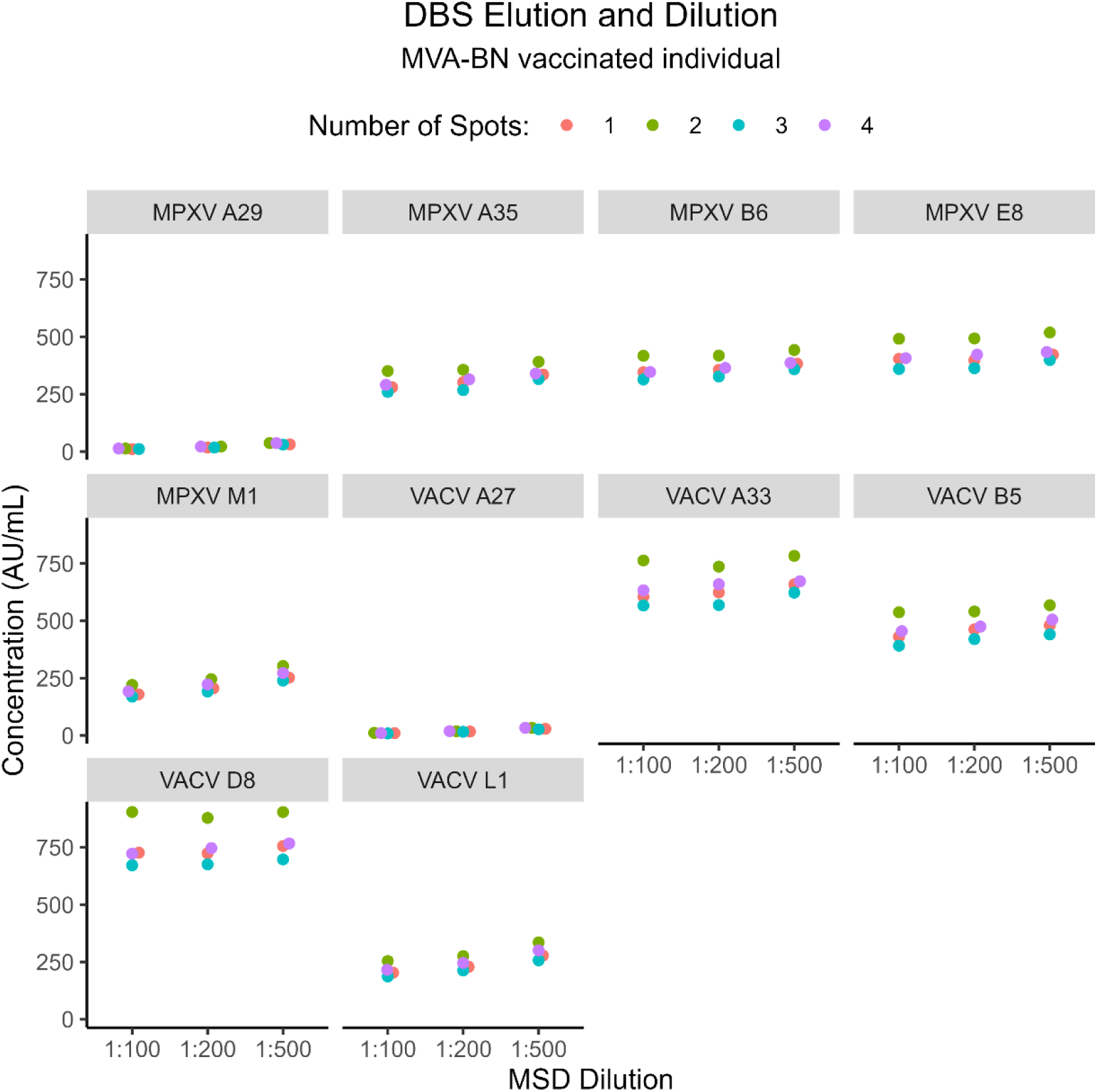
Comparison of dried blood spot elution and dilution methods within one MVA-BN vaccinated individual (with recent vaccination). Four elution methods (1 spot in 100 µL PBS, 2 spots in 150 µL PBS, 3 spots in 250 µL PBS and 4 spots in 350 µL PBS) and three Meso Scale Discovery dilutions were tested (1:100, 1:200, 1:500).

**Figure S2.**
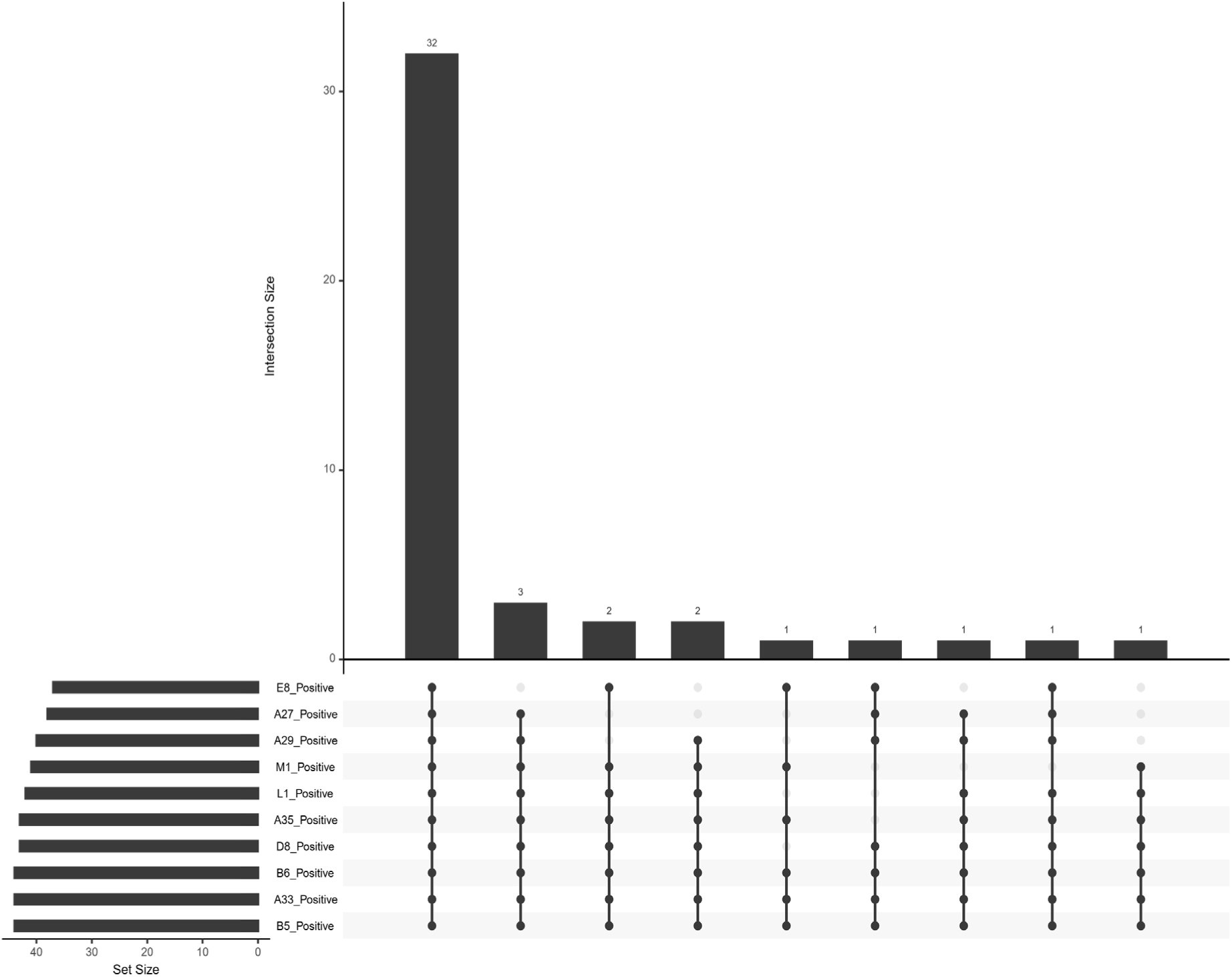
Upset plot of seroreactive samples from those with ≥ 6 individual antigenic cut-off values. Horizontal bar graphs display absolute number of samples larger than an antigen’s cut-off value. Samples that are seroreactive to multiple (or combinations of) antigens are depicted vertically. Combinations are ordered by frequency.

**Table S1.**
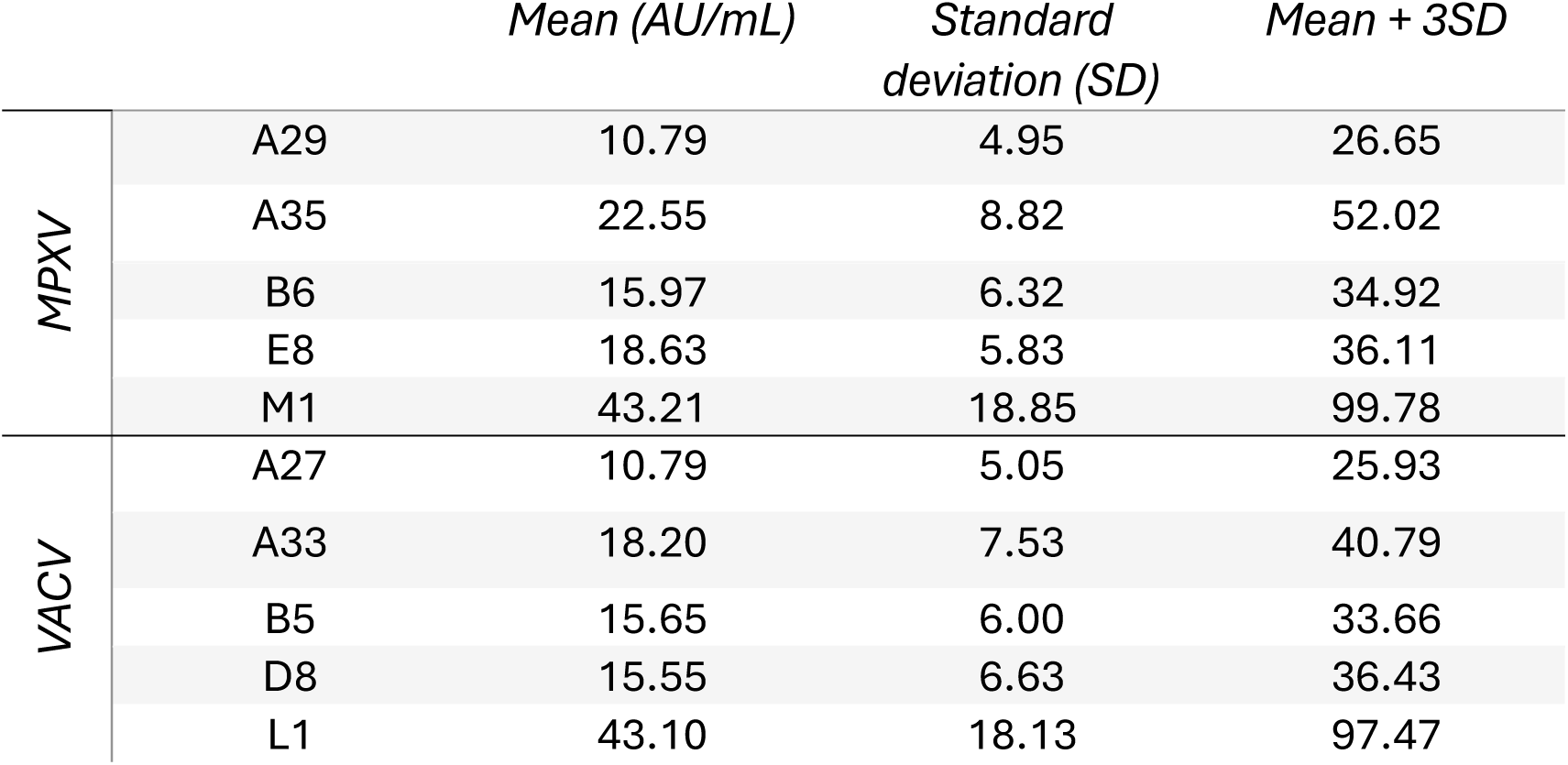
Serological cut-off values. Negative cohort derived from seven individuals (four female and three male). Median age of the negative cohort was 29 years.

**Table S2.** Number of samples seroreactive to each antigen.

|  |  | *Seroreactive* | *Unreactive* |
| --- | --- | --- | --- |
| MPXV | A29 | 58 | 145 |
|  | A35 | 51 | 152 |
|  | B6 | 55 | 148 |
|  | E8 | 56 | 147 |
|  | M1 | 54 | 149 |
| VACV | A27 | 49 | 154 |
|  | A33 | 63 | 140 |
|  | B5 | 64 | 139 |
|  | D8 | 49 | 154 |
|  | L1 | 56 | 147 |

**Table S3.** GMCconcentrations of seropositive samples.

|  |  | *GMC (95% CI)* | *GSD* |
| --- | --- | --- | --- |
| MPXV | A29 | 36.12<br>(30.74-42.45) | 1.31 |
|  | A35 | 72.03<br>(63.91-81.18) | 1.22 |
|  | B6 | 49.37<br>(44.30-55.03) | 1.20 |
|  | E8 | 47.61<br>(41.95-54.04) | 1.23 |
|  | M1 | 135.69<br>(115.94-158.80) | 1.30 |
| VACV | A27 | 31.70<br>(27.65-36.34) | 1.25 |
|  | A33 | 61.07<br>(54.00-69.10) | 1.23 |
|  | B5 | 50.03<br>(45.20-55.38) | 1.18 |
|  | D8 | 46.78<br>(41.22-53.10) | 1.23 |
|  | L1 | 134.46<br>(114.52-157.89) | 1.30 |

